# Did England’s Teenage Pregnancy Strategy reduce pregnancy rates in England? A Natural Experiment study

**DOI:** 10.1101/2020.05.12.20099002

**Authors:** Andrew J Baxter, Ruth Dundas, Frank Popham, Peter Craig

**Affiliations:** MRC/CSO Social & Public Health Sciences Unit, University of Glasgow

**Keywords:** Adolescent pregnancy, maternal health, public health, policy evaluation

## Abstract

**Objective:** To re-evaluate the impact of England’s Teenage Pregnancy Strategy (1999 to 2010) on pregnancy and birth rates. Hailed as a unique, nation-wide, comprehensive, evidence-based intervention, the strategy has been promoted as a reproducible model for other countries with high teenage pregnancy rates.

**Design:** Controlled interrupted time series and synthetic control analyses using routinely collected data on births and abortions in 16 countries.

**Setting:** The Strategy was published in July 1999 and implemented from 2000-2010, with increased investment in areas with higher rates of under-18 pregnancies from 2006 onwards.

**Participants:** Women aged under 20 living in England during the intervention period were considered to be the target population. Women in Scotland and Wales were the control population in our interrupted time series analyses. Women from European and English-speaking high-income countries were the control population in our synthetic control analyses.

**Main outcome measures:** The pregnancy rate among women aged under-18 was our primary outcome, as this was the target of the Strategy. We used under-18 births and under-20 pregnancies as secondary outcomes.

**Results:** In the controlled interrupted time series analyses, trends in rates of teenage pregnancy in England were similar to Scotland (0.08 fewer pregnancies per 1,000 women per year in England; −0.74 to 0.59) and Wales (0.14 more pregnancies per 1,000 women per year in England; −0.48 to 0.76). In synthetic control analyses, under-18 birth rates were very similar in England and the synthetic control. Under-20 pregnancy rates were marginally higher in England than in the synthetic control. Placebo testing and other sensitivity analyses supported the finding of little observable effect.

**Conclusion:** Although teenage pregnancies and births in England fell following implementation of the Teenage Pregnancy Strategy, comparisons with other countries suggest the strategy had little, if any, effect. This raises doubts about whether the strategy should be used as a model for future public health interventions in England or in other countries aiming to reduce teenage pregnancy.

The protocol for the analysis was published online at https://osf.io/tdbr8/

**SUMMARY BOX:** What is already known on this topic:

- Teenage pregnancy is associated with numerous health risks, and efforts to reduce rates of pregnancy may lead to improved health outcomes
- The Teenage Pregnancy Strategy was launched in 1999 with the goal of reducing teenage pregnancies in England
- Initial observations showed a drop in rate across the strategy period; this has been interpreted as demonstrating the success of the intervention.

What this study adds:

- Our study suggests there was little or no effect of the strategy, and that the rapid decline in rates would have occurred without the intervention, in line with similar countries.

## INTRODUCTION

Teenage pregnancy is associated with numerous health risks, both to mothers and infants. Teenage pregnancies are more likely to be unintentional than are adult pregnancies.[1] Such pregnancies are also at greater risk of health problems, such as maternal anaemia, preeclampsia, infant mortality, pre-term labour, and longer and more difficult labour.[2,3] Teenage mothers are also at greater risk than their peers of poor mental health, suicide, and substance use problems.[4]

Early pregnancy is more common among women from poorer families, single-parent households, areas of greater deprivation, and those born to teenage parents.[2,3,5] Teenagers with a previous pregnancy are up to five times more likely to experience rapid repeat pregnancies.[6] Teenage parents are more likely to face barriers to further education, employment or training, and may require greater social support for parent and child health, for positive family relationships, and income and housing support.[7,8]

It is uncertain how much becoming pregnant as a teenager contributes to these poor outcomes, or whether other socioeconomic factors may be the cause of both.[9] Other action to tackle the societal structures perpetuating these inequalities may have greater effect.[10] Nevertheless, advocates of prevention argue that reduction in rates of teenage pregnancy could improve maternal and child health and reduce health and social inequalities. [2]

A Teenage Pregnancy Strategy was introduced in England in 1999, aiming to reduce under-18s’ pregnancy rates by 50% in ten years, whilst providing support to teenage mothers.[11] In research conducted at the start of the strategy, teenage pregnancy rates were noted to be higher in England than in similar countries.[2] Yearly rates of births to women aged under 20 were more than three times higher than in France and Denmark, and more than four-times higher than in the Netherlands and Switzerland.[12] Whilst birth and pregnancy rates fell in several other European countries from 1970-1990, UK rates remained higher.[2,12]

The Strategy took a multifaceted approach to reducing rates of teenage pregnancy and addressing associated health and social problems. This involved: structured and ‘joined up’ action at national and local level to ensure coordinated, equal effectiveness in all areas; improvements in pregnancy prevention resources, including contraception access, education and media-campaigns to children and parents; and greater support for young parents to remain in education and access housing and other health support.[2,13] A mid-term review in 2005 led to significant changes in implementation, including publication of new guidance for local authorities, a redesigned media campaign, new health and education programmes, and increasing access to contraception.[11,12] The Strategy was claimed to be the first of its kind, coordinating local and national action to reduce pregnancies nationwide.[2,12]

£60m of funding was allocated for the first three years of the strategy (including £12m allocated to specific projects for young parents’ housing and childcare).[2] Expenditure on the Strategy from central government, local authorities, health authorities, other government programmes and charities, came to £167.6m by the mid-term review in 2005,[12] and reached an estimated £280m by the end of strategy activity in 2010.[14]

The Strategy was deemed a success following observations of declining pregnancy rates.[11,13,15,16] Rates of pregnancy were noted to have fallen dramatically across the period of implementation, from 47.1 pregnancies per 1,000 women aged under 18, to 22.9 per 1,000 women by 2014 - a drop of 51%.[11,17] Rates have continued to decrease, reaching 17.9 per 1,000 women by 2017,[18] an observation attributed to ongoing effects of the Strategy.[13]

The Strategy has been promoted as a unique, national approach with the cost justified by the observed fall in pregnancies.[15] It has been held up as a replicable model for ongoing national and local government action in England and for implementation in other countries.[13,19-22].

There have been two previous evaluations of the Strategy. In the first, the Teenage Pregnancy Strategy Evaluation Team measured pregnancy rates relative to the 1998 baseline rate in England and in comparison with control countries.[12] They observed a small decrease in under-18 pregnancies in England, but little difference from Scotland and Wales. They suggest that this comparison may underestimate the effects of the Strategy, due to contamination and the effects of other interventions in Scotland and Wales.[12] In comparisons with stable or increasing under-20 birth rates in other European countries, England showed small drops in rates in the first years of the Strategy. [12] A second evaluation, published in the Lancet in 2016,[17] reported a 51% drop in under-18 pregnancies from 1998 to 2014, as well as a 50% drop in under-18 births from 2004-2014. This was compared with a mean reduction of 22% in under-18 births across 28 European comparison countries (excluding Scotland and Wales).[17] The study concluded that the Strategy, "alongside other social and educational changes, has probably contributed to a substantial and accelerating decline in [under-18] conceptions”.[17]

No other research has estimated the Strategy’s effect whilst attempting to account for other causes using comparative analysis. The substantial cost of the teenage pregnancy strategy, and its promotion as a model for other countries, mean that reliable estimates of its impact are important for future policy making. We tested the effectiveness of the teenage pregnancy strategy in two ways. In our first analysis, we chose Scotland and Wales as comparators given their similarity to England in other policy, economic and cultural factors which may affect teenage pregnancy rates. We used interrupted time series methods to compare each country with England across the implementation period and up to most recent observations. To account for potential contamination among neighbouring UK countries, in our second analysis we compared birth and pregnancy rates in England with those of a wider pool of potential control countries using synthetic control methods.[23]

## METHODS

### Data collection

In each analysis, we set the intervention start as 1999. For the interrupted time series analyses, we extracted rates of teenage pregnancy directly from the Office for National Statistics report for England and Wales,[18] and Information Services Division report for Scotland,[5] for all reported age groups (under-16, under-18 and under-20). Both sources used the same calculation, summing recorded births, still births and abortions in each age group and dividing by the estimated age group female population.[5,24] Scottish rates were only reported for 1994 onwards, so to supplement these we used records of Scottish births,[25] abortions,[26] and estimates of population,[27] by age to estimate Scottish under-18 pregnancy rates from 1987 to 1993 to match the earliest data available for England and Wales. We did not include Northern Ireland due to the unreliability of estimates of abortions.[28]

We considered pregnancy to women aged under-18 as a target outcome, using comparisons with England-only data as a primary analysis.

In secondary analyses, to test using other age groups and for longer pre-intervention periods, we used England and Wales combined data as England-only data was not available. Aggregated England and Wales rates were compared with Scotland to test for effects on under-16 and under-20 pregnancies from 1992-2016 as secondary populations, and under-18s from 1987 as a secondary measure over a longer time-period. We compared England only data with England and Wales combined data for years recording both to assess the suitability of the combined data as a proxy for exposed England. England contributed around 95% to both population and pregnancy outcomes and rates were very similar across all years, suggesting that aggregated England and Wales rates were a good indicator in the absence of England-only data.

For the synthetic control analyses, we selected countries for comparison based on cultural, political, geographical and economic similarity to England. We sought data on teenage births and pregnancies for all Euro-peristat nations,[29] and other high-income Anglophone countries. We aimed to collect data recording births and pregnancies for at least eight time points before and after the intervention.

We used data estimating births by age of mother from the Human Fertility Database,[30] populations from the Human Mortality Database,[27] and numbers of abortions to women under-20 from the WHO Health for All Explorer.[31] Data on births, abortions and pregnancies for countries not included in the Human Fertility Database were sought from national statistics websites. Pregnancy and birth rates for the USA were extracted from the Guttmacher Institute report. These were calculated using population, birth and abortion data from the National Centre for Health Statistics and the Center for Disease Control.[32] Pregnancy and birth rates for New Zealand were calculated from Statistics New Zealand reports on births and abortions, combined with Human Mortality Database population estimates.[33,34] Full details are given in Supplementary File Section A.

Four countries were excluded for which no data or incomplete data were available (Austria, Australia, Ireland, and Canada). This was due to different age groupings, insufficient time points or no reliable records of abortions. Finally, we excluded eight European countries that were either in Yugoslavia or the USSR, or were USSR-backed, as they had turbulent histories around this time making them less useful as comparators (Hungary, Estonia, Lithuania, Slovenia, Czechia, Poland, Croatia, and Bulgaria). The final selection of fifteen control countries is shown in Box 1.

#### Box 1 Countries selected to construct synthetic controls

|  |  |
| --- | --- |
| Denmark | Norway |
| Finland | Portugal |
| France | Scotland |
| Germany | Spain |
| Iceland | Sweden |
| Italy | Switzerland |
| Netherlands | USA |
| New Zealand |  |

Outcome rates for 1990-2013 were calculated as the earliest and latest dates with data available for a sufficient set of comparison countries. We calculated under-18 birth rates by summing all births to women aged under-18 and dividing by total populations aged 15-17, matching the age group reported by ONS and ISD Scotland.[5,24] We used births only as we did not find reliable data estimating abortions to under-18s for all countries, and so we could not estimate total pregnancies. We calculated under-20 pregnancy rates by summing all under-20 births, adding total abortions to women under 20 and dividing by total populations aged 15-19. We recalculated England and Wales’ and Scotland’s under-18 birth-rates from these datasets to make them comparable. England and Wales were used as a single unit as only combined data were available.

Estimates of yearly gross domestic product (GDP), mobile phone ownership, proportion of females in population and proportion of population resident in urban settings for years 1990 to 2013 were extracted from World Bank open data as predictor variables for the synthetic control models.[35] Public spending on education as a proportion of GDP for the years 1990 to 2013 was extracted from OECD data.[36]

### Statistical analysis

All analyses used R (v 3.6.1),[37] and RStudio.[38] We built a Shiny app to carry out the ITS analysis.[39] All R packages used are listed in Supplementary File Section A.

To compare England with Scotland and Wales we used interrupted time series methods.[40-42] In our preparatory models we fitted a trend line to England observations before the start of the strategy in 1999 to estimate the baseline trend as an hypothesis of the trajectory England would have followed in the absence of the strategy.

We then fitted an intervention trend line to data from 1999 to 2016 to estimate the changes in trend and level from the start of the strategy. This allowed years beyond the 2010 end of the intervention to contribute to estimates of its effects, consistent with previous evaluations. We visually inspected the pregnancy rates across this period to determine if any changes immediately after the 2010 end indicated a temporary effect of the strategy, requiring exclusion of later data. This trend remained consistent and so these time points were used in all analyses as assumed ongoing effects of the strategy.

Our comparison models used Scotland and Wales as control populations to estimate the expected changes at 1999 in the absence of the strategy. Changes in level and trend seen in Scotland and Wales were subtracted from those seen in England to give estimates of the strategy’s effects, corrected for background changes common to all three countries.

To improve the fit of the pre-intervention rates, we added a ‘pill scare’ dummy variable across all three countries for all dates from 1996 onwards. This aimed to account for the hypothesised effects of a warning issued concerning the safety of oral contraceptive pills in 1995 and the subsequent fall in contraceptive use.[12,17,43]

Inspection of pre-intervention trends between England and controls indicated that all three countries closely followed the same pattern before the strategy. Therefore, the primary model used the assumption of pre-intervention parallel trends, allowing more stable predictions from the limited pre-intervention data. After examining rates across all three countries, we saw a similar trend change occurring from 2008 onwards, dividing the post-intervention period into two segments. In sensitivity analyses we treated 2008 as a common shock across all countries and allowed a common trend change to better fit the observations. T o test whether allowing for a phase-in period improved model fit, we excluded data for the years immediately following the start of the intervention. This made no difference to fit or prediction, so all data were retained in final analyses.

Data for England alone was only available for 1992 onwards, giving seven pre-intervention time points. To test model sensitivity by examining longer pre-intervention time periods, we used combined England and Wales data, available from 1987, to compare with Scotland.

We tested for autocorrelation using Durbin-Watson tests, and autocorrelation and partial-autocorrelation function plots. We applied corrections to our final models when autocorrelation was evident across all three tests. Finally, we extracted coefficients and 95% confidence intervals for difference in level and trend change seen in England over controls at each time period and used these as markers of change due to intervention.

In our second analysis, we used synthetic control methods to construct a comparison unit from a weighted average of other countries’ rates, fitted to pre-intervention England and Wales observations. We used under-18 birth rates as a primary outcome and under-20 pregnancy rates as a secondary measure to get a clearer estimate of effect on pregnancies rather than births. Initial models used each country’s mean rate across the whole preintervention period (1990-1998) as a single predictor to construct the synthetic England. To improve the pre-intervention control fit, we used a data-driven approach by finding optimal groupings of years and calculating means for each period as a predictor, to account for the non-linear pattern of the yearly rate changes. The optimal grouping was chosen as a combination of as few groups as possible and a minimised mean squared prediction error. After selecting the best pre-intervention fit rate-only model, we tested the effects of adding other predictors on the overall model fit.

To test our models, we conducted several robustness checks and sensitivity analyses. Removing England and Wales data, we repeated the synthetic control analyses for each of the other countries as placebos and recorded observed and predicted values. Yearly differences between observations for England and Wales and their synthetic control were plotted alongside corresponding differences calculated for the other countries and their synthetic controls to check whether England and Wales was a comparative outlier. We excluded countries with greater than 5-times the pre-intervention MSPE of England and Wales for to compare the exposed population with similarly well fit placebos. Using all comparison countries, we calculated post/pre-MSPE ratios for each country and examined their distribution to check whether England and Wales saw a large deviation from predicted post-intervention rates compared to unexposed countries. Finally, we constructed plots of observed and synthetic rates for models fitted to dummy intervention dates across 1995-1998 to examine whether the model was robust to shocks in pre-intervention years.

We performed sensitivity analyses to test the reliability of our models. We re-ran models with countries removed from the donor list to test for over-reliance on a few countries’ data. We iteratively removed the top-weighted country in each analysis, plotting yearly differences between England and Wales and the new synthetic control, and extracting pre-intervention MSPE for each to test whether results remained consistent as donor countries were removed.

## RESULTS

### Comparing England with Scotland and Wales using Interrupted Time Series methods

England saw a 60% drop in under-18 pregnancies between 1998 and 2016, from 46.6 to 18.8 pregnancies per 1,000 women (Figure 1). Across the same period, Scotland saw a reduction in pregnancies of 58% (from 44.7 to 18.9 pregnancies per 1,000 women) and Wales of 62% (from 55.0 to 20.9 pregnancies per 1,000 women). All three countries saw a small jump in rates in 1996, consistent with hypothesised effects of the 1995 ‘pill scare’ leading to less contraceptive use.[43]

**Figure 1.**
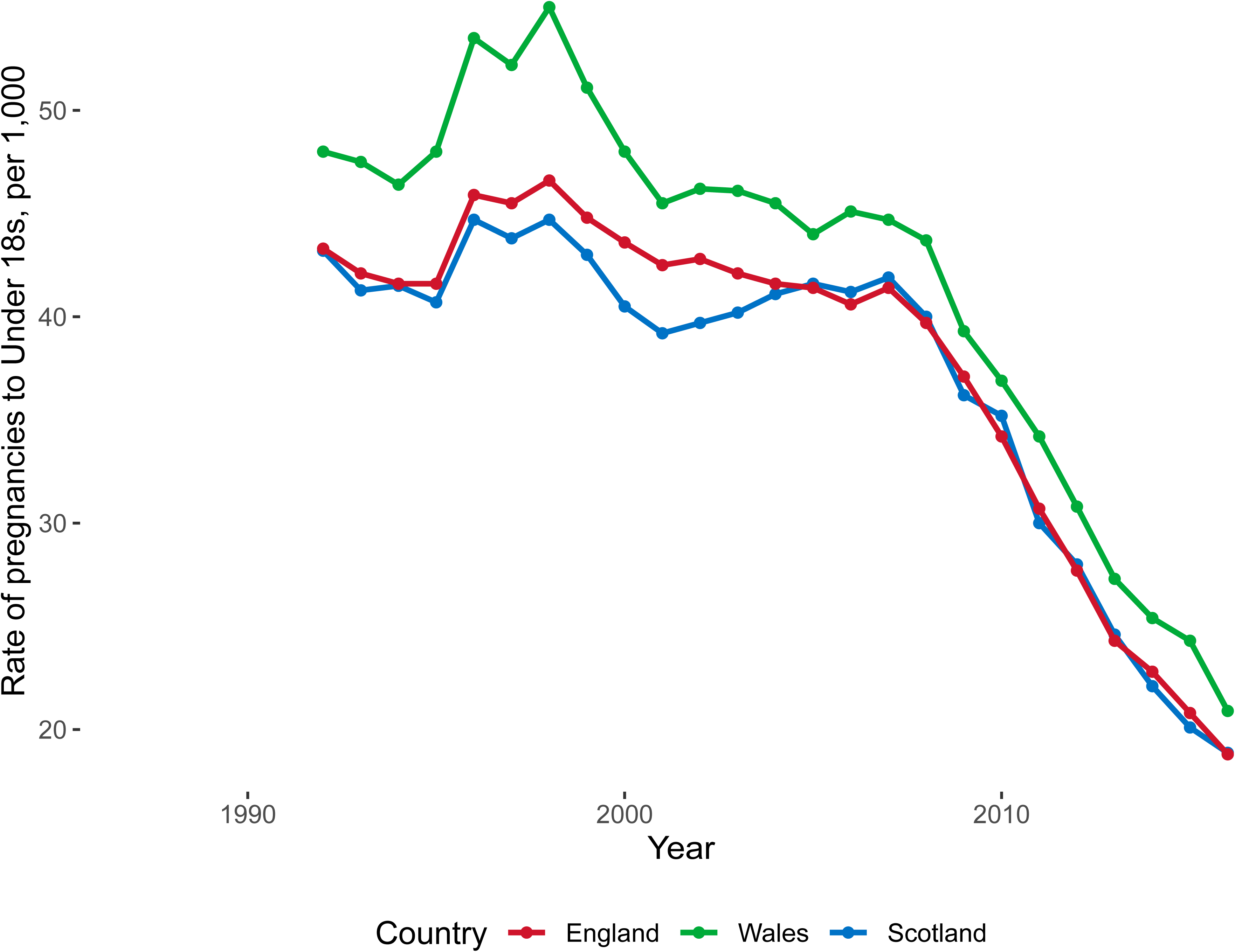
Under-18 pregnancy rates across England, Wales and Scotland 1992-2016.

Figure 2a from an interrupted time series (ITS) analysis using England-only before and after comparison shows an initial upward trend of 0.70 more pregnancies per year per 1,000 women (95%CI: −0.34 to 1.74) during the pre-intervention period that is reversed by a clear change in trend from 1999 onwards, with an accumulating 2.22 fewer pregnancies per 1,000 women per year than predicted from pre-strategy rates (95%CI: −3.49 to −0.95). Addition of a pre-intervention change in level that accounts for the ‘pill-scare’ in 1996 improved model fit for the pre-intervention period (Figure 2b). The corrected pre-intervention trend was −0.11 per year (95%CI: −1.10 to 0.88), with a reduction in trend from 1999 onwards of an additional accumulating 1.41 fewer pregnancies per 1,000 women per year than predicted (95%CI: - 2.58 to −0.24; Figure 2b). The 1996 corrector was used in further analyses. No significant level changes were observed at 1999.

**Figure 2.**
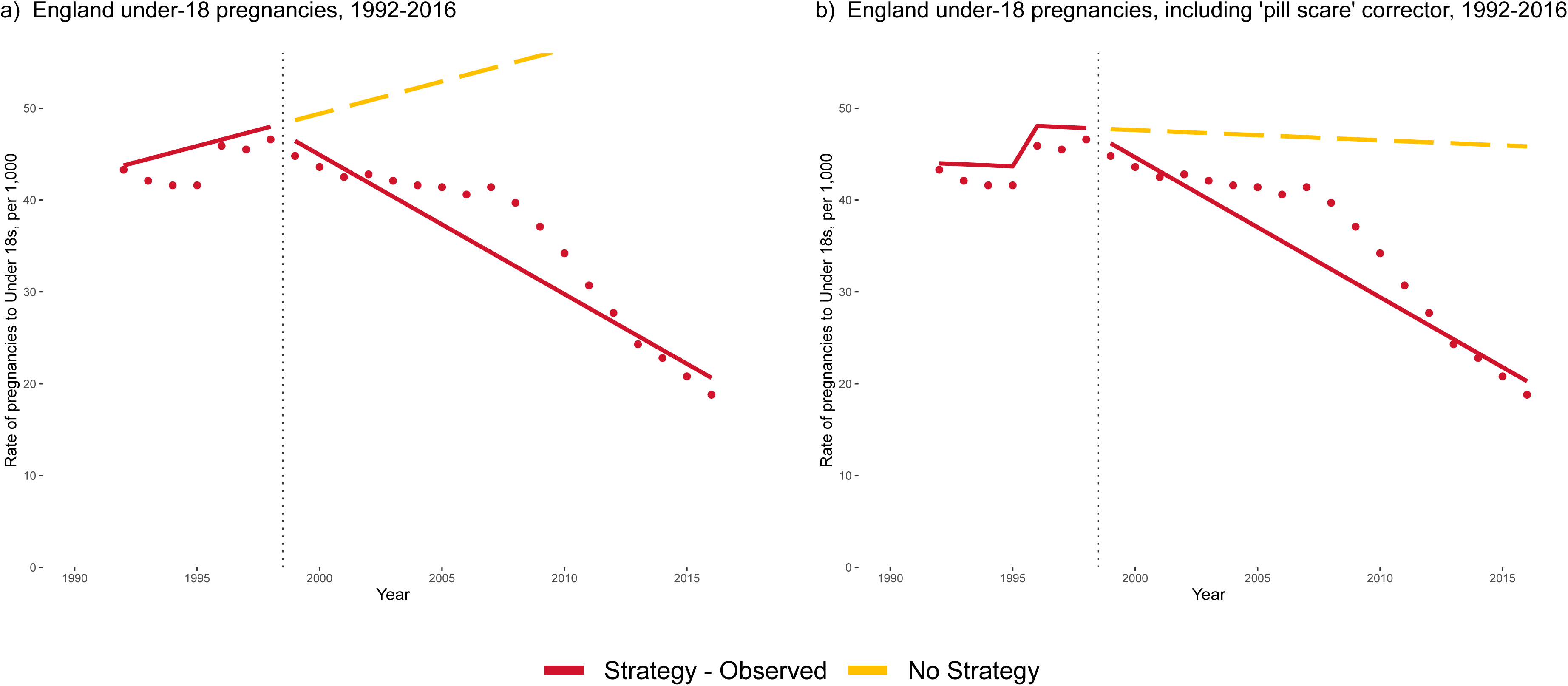
Uncontrolled interrupted time series comparisons of England’s before and after under-18 pregnancy rates. In the initial comparison without corrector in a), England saw a level change of −0.03 (−3.08 to 3.01) at 1999 and a trend change of −2.22 (−3.49 to −0.95). With the addition of a corrector at 1996 in figure b), the level change became −1.14 (−2.62 to 2.34) and the trend change −1.42 (−2.58 to −0.24). All models are corrected for autoregression at lag 1.

In the controlled ITS analyses, these effect sizes were greatly decreased. Level and trend changes in Scotland and Wales data were applied to England’s pre-intervention trend to predict a ‘No Strategy’ control, assuming that the observed changes in control countries would have occurred in England without the TPS. In comparison with a control constructed from Scotland’s level and trend changes, there was a decrease of 0.08 pregnancies per 1,000 women per year in England (95%CI: −0.74 to 0.59; Figure 3a). In comparison with Wales, England saw a small increase over control of 0.14 pregnancies per 1,000 women per year (95%CI: −0.48 to 0.76; Figure 3b). All controlled models showed results consistent with a null effect of the Teenage Pregnancy Strategy.

**Figure 3.**
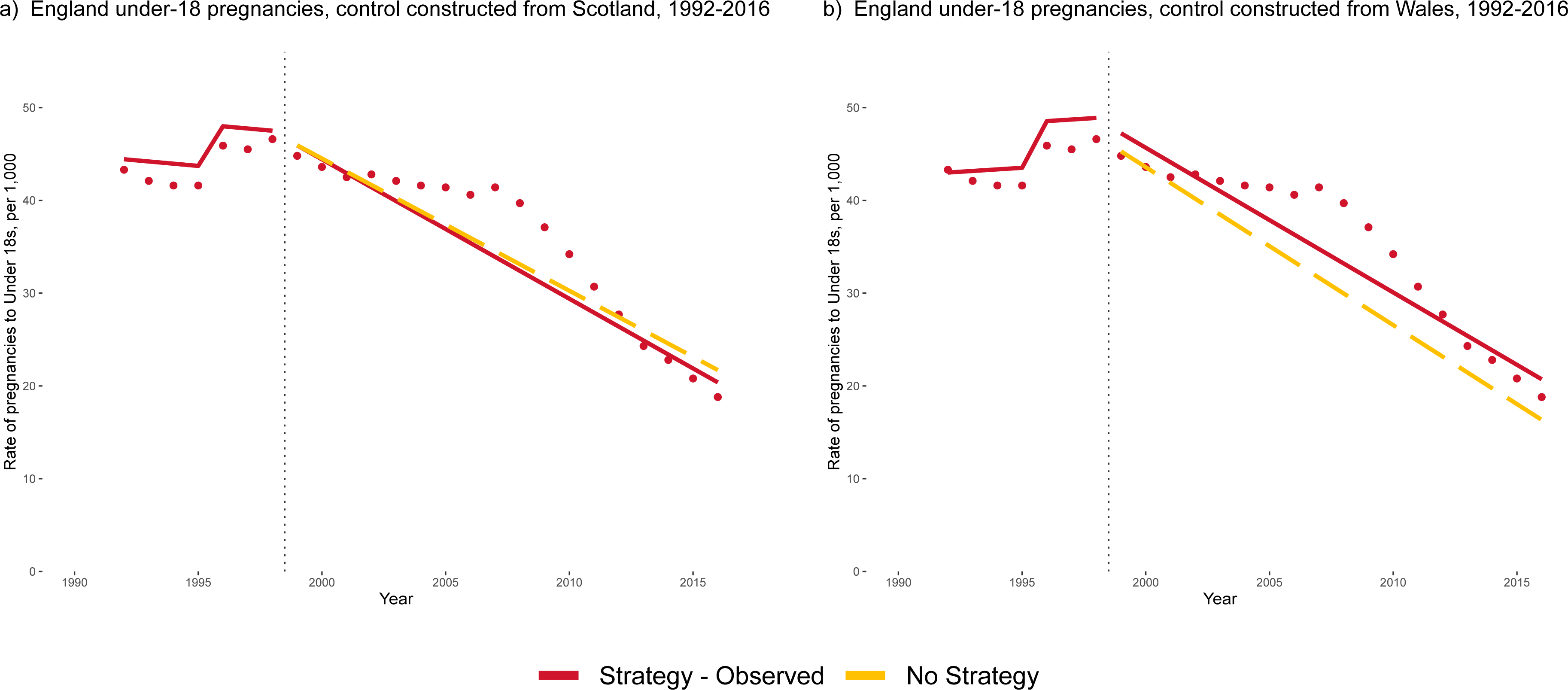
Controlled interrupted time series comparisons, using data from Scotland and Wales to predict England’s changes in rates without the TPS. In comparison to control adjusted to match Scotland’s change in level and trend at 1999, England saw a level change of 0.06 (−4.03 to 4.16) and a trend change of −0.08 (−0.74 to 0.59; graph a). In comparison to control adjusted using Wales’ data, England saw a level change of 1.81 (−2.30 to 5.91) and a trend change of 0.14 (−0.48 to 0.76; graph b). All models are corrected for autoregression at lag 1.

In a further set of analyses (Supplementary File Section B), we allowed for a ‘common shock’ at 2008 to account for a common change in trend in all three countries from 2008 onwards, but these also revealed no significant differences between England and control. Finally, we combined England and Wales data to examine longer pre-intervention periods as well as under-16 and under-20 pregnancy rates. No significant differences were seen at 1999 across these analyses.

### Comparing England and Wales with other countries using Synthetic Control methods

Our primary synthetic control model used under-18 birth rates from 15 countries and calculated means of four groupings of pre-intervention years as predictors (1990-1993, 1994, 1995, 1996-1998). We were able to construct good-fit synthetic controls to compare with England and Wales using only pre-intervention birth rates. The prediction error of this model was 0.31 births per 1,000 women per year around a mean of 16.2 births per 1,000 women across 9 years (Mean Squared Prediction Error, MSPE: 0.10; Figure 4a). This model was used as our primary comparison. The synthetic control for England and Wales was constructed from a weighted mean of Scotland (weighted 67.2%), Portugal (29.5%), the U.S.A. (1.6%) and New Zealand (1.2%). Birth rates for the synthetic control closely followed the observed birth rates in England and Wales across the whole post-intervention period. While England and Wales saw a drop in birth rates of 53% between 1998 and 2013, the control saw a 50% drop.

**Figure 4.**
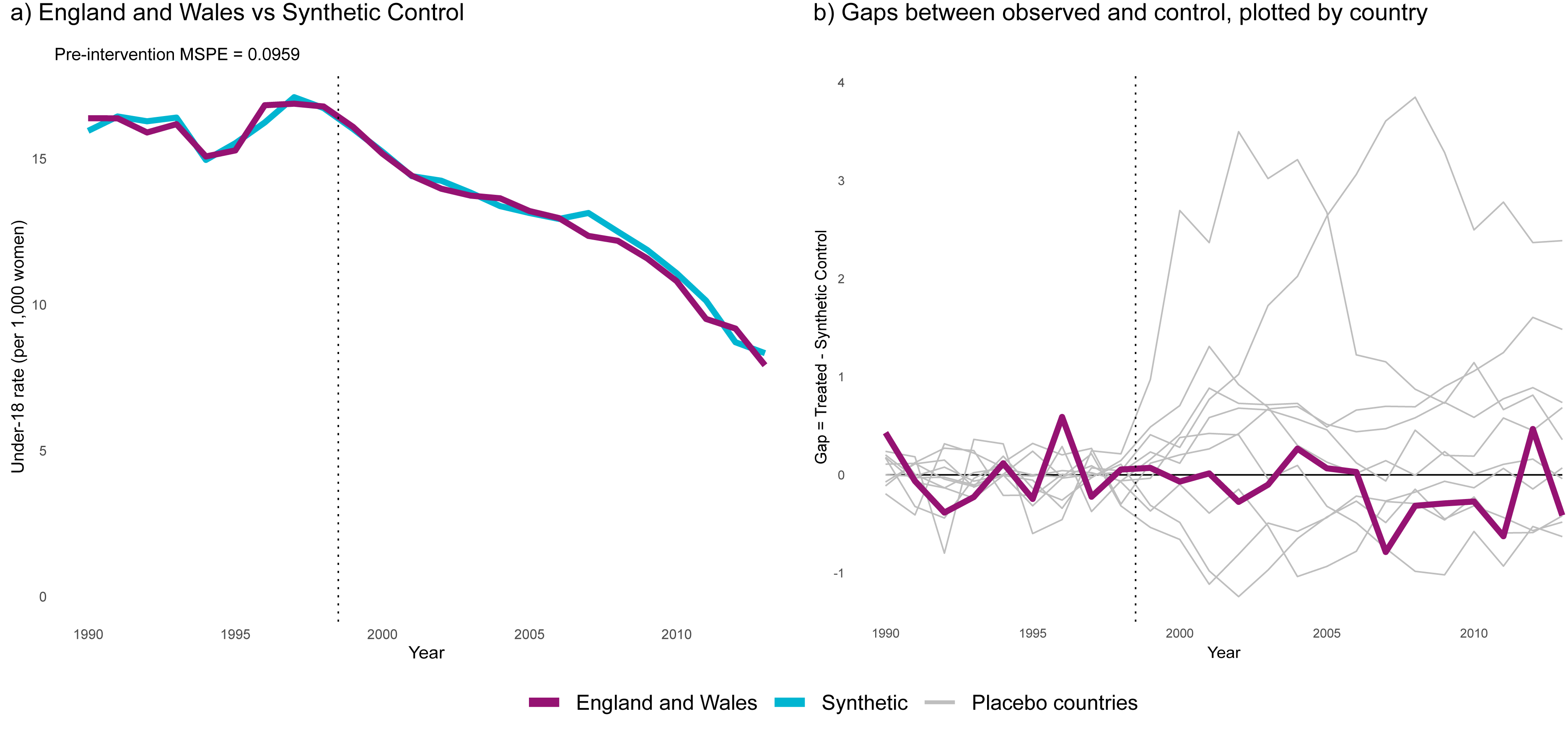
England and Wales’ observed combined under-18 birth rates compared with synthetic control, 1990-2013. In graph a, rates are plotted for pre- and post-intervention periods, with a pre-intervention fit (Mean Squared Prediction Error) of 0.10. In graph b, yearly differences between England and Wales and its synthetic control are plotted alongside similarly calculated gaps for ten placebo countries, with pre-intervention fits close to England and Wales (less than 5 times England and Wales’ pre-intervention MSPE).

Gaps between the observed rates for each country and the predicted rate for its synthetic control are plotted in Figure 4b. Post-intervention effect sizes for England and Wales fall within the range of gaps for other countries with a well-fitting synthetic control. The post/pre-MSPE ratio for England and Wales, measuring comparative variance between fitting and predicting periods, was calculated as 1.27. 13 of the 15 control countries saw a larger post/pre-MSPE ratio, indicating that the probability of observing a ratio at least this large in the absence of an effect is p = 0.88. These results are consistent with a null effect of the TPS.

Using under-20 pregnancies as a secondary outcome resulted in a slightly poorer preintervention fit, with a pre-intervention average prediction error of 2.07 pregnancies per year around a mean of 62.3 pregnancies per 1,000 women (MSPE: 4.27). Poorest fit was seen across the years 1996-1998, immediately preceding the strategy and correlated with the pill-scare jump occurring predominantly in the UK. The control saw a slightly greater decrease in pregnancy rates than England and Wales during the strategy period, but a relatively small post/pre-MSPE ratio compared to placebo countries (9.1; rank 11 out of 16 countries; p = 0.68; see Supplementary File Section C). Our time-placebo analyses tested the model with dummy interventions across 1995-1998. When set at 1995 and 1996, the predicted control rates were much lower than England and Wales, but 1997 and 1998 produced controlled models very similar to the true model. These results are consistent with a null effect of the strategy.

All sensitivity tests are reported in Supplementary File Sections C and D. When we removed Scotland from the donor pool, we saw poorer pre-intervention fit and a small drop in under-18 birth rates in England and Wales compared to control throughout the strategy-period. This difference from control was still relatively small compared to the noise seen in placebo countries and gave no strong indication of an intervention effect. Optimising model-fit to the immediate pre-intervention years 1996-1998 to account for the ‘pill scare’ did not produce an effect. Across all other analyses, we saw poorer predictor fit than our primary and secondary models, and consistent, small gaps between England and Wales and control, with higher birth and pregnancy rates in England and Wales across the intervention period. This is consistent with a null effect of the strategy.

## DISCUSSION

### Main findings

We find no evidence of an effect of the Teenage Pregnancy Strategy on rates of teenage pregnancies or births in England between 1999 and 2016. Analysis of England-only data showed a clear change in trend during the Strategy period, consistent with previous observations.[17] However, the similar changes observed in other UK, European and English speaking countries suggest that England may have seen a similar fall in teenage pregnancy in the absence of the Strategy. This finding of little, if any, impact was consistent across two methods using different datasets, and was robust to sensitivity analyses.

### Strengths and limitations

We used publicly available, reliable data from several sources which was comparable across countries. Whilst natural experiment methods each have weaknesses which threaten the confidence of causal inference,[40] our use of two methods and several comparisons sought to account for these. The coherence of conclusions reached through all analyses strengthens our findings.

In ITS models, data were limited in terms of periods of observation for each age group. Under-18 pregnancy rates for England alone represented our primary outcome; however, in published data, these were only available for seven pre-intervention time points.[18] A minimum of eight time points for ITS analyses are usually recommended; our primary models may have lacked power to detect small changes.[44-46] Sensitivity analyses using England and Wales data with more pre-intervention time points were used to account for this and achieved consistent results.

The outcome measures for each analysis had several limitations. Rates calculated for the UK using the ONS methods of adding births, still births and recorded abortions are not able to account for miscarriages and illegal abortions.[24] As all three countries had similar laws, healthcare and access to abortion clinics, we judged that these errors would be unlikely to have been differentially distributed across countries and therefore would produce negligible bias in comparative analyses. In comparisons with countries outside of the UK, we used counts of births and sums of births and abortions to estimate pregnancies. These data were not able to be corrected in the same manner as ONS and ISD Scotland data, and so are less reliable measures of actual pregnancy rates. However, they provided estimates of births and pregnancies to teenage mothers using consistent definitions and data sources, which were comparable across countries.

In SC analyses, under-18 pregnancy rates were not directly calculable as under-18 abortion estimates were not reliably available in a consistent way across all countries. The two measures, under-18 births in our primary analyses and under-20 pregnancies in sensitivity analyses, were used in place of under-18 pregnancies and gave consistent results.

In our SC sensitivity tests, pre-intervention fit was poorest across the period 1996-1998, particularly after removal of Scotland. This increase in rates, observed mainly in UK countries (and across all measures used) has been attributed to media messages surrounding suggested health risks of certain contraceptive pills around 1995 – the ‘pill scare’.[2,12] The event was confined to the UK and was followed by reductions in oral contraceptive use.[43] This may have contributed to the higher rates of pregnancy than control across the whole period from 1995-2013 and may explain the time-placebo test results showing large differences from 1995 and 1996 dummy intervention dates. However, when we accounted for this by optimising the pre-intervention fit to the years 1996-1998 alone, we still saw no difference from control that would be consistent with an effect of the Strategy.

Concerns have been raised about using Scotland and Wales as comparators to identify the effects of the English strategy, either because they may have been contaminated by the media campaign,[47,48] or because they implemented similar policies.[12,17] Contamination is a possibility, but any spill over effects should be weaker than the effect of direct exposure to the strategy. Our analyses would have been able to detect any additional effect in England associated with full exposure to the strategy, consistent with an expected dose-response effect of more intense action and focus on England. An alternative hypothesis is that the strategy’s media campaign was predominantly responsible for the very similar observed changes across England, Wales and Scotland, and that other elements of the Strategy had little or no effect. However, Wellings et al.[17] report differential effects associated with strategy spending between local authorities in England. Such effects should also be evident in cross-border differences yet the trends in England, Scotland and Wales are all very similar. Evaluations of other teenage pregnancy interventions implemented in Scotland do not suggest effects that could mask a substantial effect of the English strategy.[49,50] Contamination and spill over effects should not affect the validity of the synthetic control analyses.

Our analysis was restricted to evaluating the aim of reducing teenage pregnancy, whilst providing support to young mothers was an additional strategy aim.[2] As Lawlor, Shaw and Johns argue,[10] such aims may have positive effects on the health and social inequalities associated with teenage pregnancy. The strategy may have been effective for these outcomes.

### Implications

The lack of detectable differences between England and controls is consistent with the hypothesis that the Teenage Pregnancy Strategy had minimal or no effect on pregnancy rates. The strategy had several aims,[2] and whilst our study did not look at outcomes related to support for young mothers and infants, rates of pregnancy were considered a key measure of progress of the strategy overall.[11]

Despite the large drops after 1999, teenage pregnancy and birth rates in England remain comparatively high amongst the countries considered here. Teenage pregnancy and birth rates remain a target of policy, and current policy cites the strategy as a model.[19] Our findings suggest the Teenage Pregnancy Strategy should not be relied upon as a means of further reducing pregnancy rates in England, or as a replicable model for other countries with high pregnancy rates.

It is not yet clear what produced the observed changes. Further research could test other hypothesised causes behind the observed rates across several countries during the time-period. Other potential causes have been suggested, such as economic changes, improvements in contraception technologies, changes in other social welfare policies and greater access of young women to education.[51-53] There are also suggestions the that introduction of smartphones to society from 2007 onwards may have contributed to global trends in decreasing adolescent sexual activity.[54] This is consistent with the observed common change in pregnancy trends at 2008 across England, Wales and Scotland (Supplementary File Section B). These changes are likely to have influenced rates across several countries. These causes may inform future policy development by highlighting new modifiable causes or opportunities for effective intervention.

## CONCLUSIONS

We found no evidence of any impact of the Teenage Pregnancy Strategy on rates of pregnancy or birth among adolescents in England. Our analyses suggest that the same pattern of decreasing rates would have occurred without the strategy. The strategy should not be used as a model for future public health interventions in England or in other countries.

## Data Availability

All code used for data cleaning and analysis is available online at https://github.com/andrewbaxter439/teen-preg-project (archived at https://dx.doi.org/10.5281/zenodo.3761903) and https://github.com/andrewbaxter439/ITS_shinyapp (archived at https://dx.doi.org/10.5281/zenodo.3690804). Data sets of calculated outcomes and predictors for countries are included; raw data used to calculate these is available from respective websites, detailed at these repositories.

https://osf.io/8u9jp/

https://github.com/andrewbaxter439/ITS_shinyapp

https://github.com/andrewbaxter439/teen-preg-project

## Supplementary File

Section A - Data sources and code used in analyses

Section B - Interrupted time series analysis outputs

Section C - Synthetic control analysis outputs

Section D - Synthetic control iterating over year groupings and removing control countries

## Declarations

### Authors’ contributions

RD, FP and PC proposed that the evaluation be done using natural experiment methods. AB selected the methods, retrieved necessary data, and constructed the R code to carry out analyses and output results and visualisations. AB drafted the manuscript and all authors contributed and approved the final submitted version.

### Ethical approval

Ethical approval was not required

### Patient and public involvement

No patients or members of the public were involved in the design and conduct of this study. Dissemination of findings to participants was not applicable.

### Transparency statement

AB affirms that the manuscript is an honest, accurate, and transparent account of the study being reported; that no important aspects of the study have been omitted; and that all discrepancies from the study as originally planned have been explained.

## Acknowledgements

**Code and data for reproducibility**

All code used for data cleaning and analysis is available online at https://github.com/andrewbaxter439/teen-preg-project (archived at https://dx.doi.org/10.5281/zenodo.3822193) and https://github.com/andrewbaxter439/ITS shinyapp (archived at https://dx.doi.org/10.5281/zenodo.3822198). Data sets of calculated outcomes and predictors for countries are included; raw data used to calculate these is available from respective websites, detailed above.

**Open access**

This is an open access article distributed in accordance with the terms of the Creative Commons Attribution (CC BY 4.0) license, which permits others to distribute, remix, adapt and build upon this work, for commercial use, provided the original work is properly cited. See: http://creativecommons.org/licenses/by/4.0.

